# Pilot study of the association between microbiome and the development of adverse posttraumatic neuropsychiatric sequelae after traumatic stress exposure

**DOI:** 10.1101/2023.03.01.23286577

**Authors:** Abigail L Zeamer, Marie-Claire Salive, Xinming An, Stacey L House, Francesca L Beaudoin, Jennifer S Stevens, Donglin Zeng, Thomas C Neylan, Gari D Clifford, Sarah D Linnstaedt, Scott L Rauch, Alan B Storrow, Christopher Lewandowski, Paul I Musey, Phyllis L Hendry, Sophia Sheikh, Christopher W Jones, Brittany E Punches, Robert A Swor, Lauren A Hudak, Jose L Pascual, Mark J Seamon, Erica Harris, Claire Pearson, David A Peak, Roland C Merchant, Robert M Domeier, Niels K Rathlev, Brian J O’Neil, Paulina Sergot, Leon D Sanchez, Steven E Bruce, Ronald C Kessler, Karestan C Koenen, Samuel A McLean, Vanni Bucci, John P Haran

## Abstract

**Background:** Patients exposed to trauma often experience high rates of adverse post-traumatic neuropsychiatric sequelae (APNS). The biologic mechanisms promoting APNS are currently unknown, but the microbiota-gut-brain axis offers an avenue to understanding mechanisms as well as possibilities for intervention. Microbiome composition at the time of trauma exposure has been poorly examined regarding neuropsychiatric outcomes. We aimed to determine whether baseline the gut microbiomes of trauma-exposed emergency department patients who later develop APNS have dysfunctional gut microbiome profiles and discover potential associated mechanisms.

**Methods:** We performed metagenomic analysis on stool samples (n=51) from a subset of adults enrolled in the Advancing Understanding of RecOvery afteR traumA (AURORA) study. Twelve-week post-trauma outcomes for post-traumatic stress disorder (PTSD) (PTSD checklist for DSM-5), normalized depression scores (PROMIS Depression Short Form 8b) and somatic symptom counts were collected. Generalized linear models were created for each outcome using microbial abundances and relevant demographics. Mixed-effect random forest machine learning models were used to identify associations between APNS outcomes and microbial features and metabolic pathways.

**Results:** Microbial species, including *Flavonifactor plautti* and *Ruminococcus gnavus,* which are associated with inflammation and poor health outcomes, were found to be important in predicting worse APNS outcomes. Notably, worse APNS outcomes were highly predicted by decreased L-arginine related pathway genes and increased citrulline and ornithine pathways.

**Conclusions:** Pro-inflammatory microbes that are enriched in individuals who develop APNS. More notably, we identified a biological mechanism through which the gut microbiome reduces global arginine bioavailability, which also has been demonstrated in patients with PTSD.

## Introduction

Adverse posttraumatic neuropsychiatric sequelae (APNS) such as posttraumatic stress disorder (PTSD), depression, and somatic symptoms are common after traumatic stress exposure (1–6). Contemporary limitations in understanding the pathogenesis of APNS are a barrier to developing effective primary and secondary preventive interventions(1, 7). Novel multidisciplinary approaches applying methodologic advances from other areas of neuroscience to the early post-trauma period may advance understanding of APNS pathogenesis.

One such area is the study of the microbiome-gut-brain axis. The microbiome-gut-brain axis has been demonstrated to have an important influence on brain function in neuropsychiatric disorders(8–10), and has been hypothesized to play an important role in trauma-related neuropsychiatric disorders(11). Gut microbes affect central nervous system function through the production of metabolites and neurochemicals(10). For example, short-chain fatty acids such as butyrate support gut homeostasis by maintaining gut barrier integrity and inducing anti-inflammatory factors which cross the blood-brain barrier (12, 13). In contrast, *Enterobacteriaceae* expansion can augment neuroinflammatory processes through the lipopolysaccharide (LPS) endotoxin-mediated activation of the Toll-like receptor 4 (TLR4) pathway, leading to the production of pro-inflammatory cytokines(14).

While specific mechanisms responsible for it remain incompletely understood, dysregulated immunity and elevated inflammation are risk factors for PTSD(15). Given the above known influence of the gut-brain axis on neuroinflammation, these data support the hypothesis that variations in microbiome characteristics could influence APNS pathogenesis. This hypothesis is also supported by several small studies which identified broad phylum-level differences in microbiome characteristics among individuals with PTSD (n=18) as compared to those without PTSD (n=12) (16), and a study in veterans with cirrhosis which found associations between lower microbial diversity and a higher abundance of opportunistic microbes with PTSD symptoms and impaired cognition(17). This hypothesis is also supported by animal studies in which trauma was found to cause alterations in the microbiome and lead to increased local and systemic inflammation(8).

In this nested study, we explored the association between the gut microbiome and APNS development in a subsample of individuals recruited from emergency departments (ED) in the immediate aftermath of trauma as part of the AURORA study (n=74) and who also provided stool samples(1). We employed shotgun metagenomic sequencing of stool samples from patients evaluated and discharged from the ED and machine learning (ML) predictive analytics to test the hypothesis that variability in microbiome species and pro- and anti-inflammatory metabolic pathways is associated with APNS outcomes.

## Methods

### Study Population and Sample Collection

The AURORA study began in September 2017; we started enrollment into this nested study from November 2020 until February 2021. AURORA participants who completed their first outpatient remote assessment and had not received antibiotics within the previous 6 months were approached for enrollment into this nested study. Participants also could not have had any other influences that could cause major microbiome perturbations (e.g., infections, significant dietary changes). Of the 2,097 AURORA participants subjects approached, 106 agreed to participate, and we received stool samples from n=74 of these individuals (69.8%) for this nested study. Stool samples were self-collected at home by participants using OMNIgene•GUT collection kits (DNAgenotek, catalog no. OMR-200) and sent to the study laboratory. As we were recruiting for this study and collecting samples during the initial phases of the COVID-19 pandemic, immediate sample collection soon after the traumatic event that was the primary inclusion criterion for AURORA was not possible. All stool samples were self-collected at least five days after the initial ED presentation and up to 182 days. Upon receipt, samples were stored at -80°C until DNA extraction and sequencing were performed. This study was approved by the University of North Carolina’s institutional review board (IRB protocol #17-0703) and approved by the UMass Chan Medical School’s institutional review board.

### Outcome Measurements

After receiving written informed consent from eligible patients, study coordinators from each participating ED performed data collection for ED-based assessments, including baseline questionnaires. Follow-up evaluations were internet-based at the two, eight, and twelve-week intervals. We assessed posttraumatic stress symptoms using the Post Traumatic Stress Disorder (PTSD) checklist for DSM-5 (PCL-5)(18–21). Per this instrument’s guidelines, a PTSD diagnosis can be made by summing the self-reported responses to the PCL-5. We used total sum of 31 for a positive diagnosis of PTSD(22). A normalized t-score for depression severity and the diagnosis was assessed using the PROMIS Depression Short Form 8b (23). A t-score of 60 is one standard deviation lower than average and was used to designate a depression diagnosis. The Rivermead Post-concussive questionnaire (RPQ) was used to obtain a count of somatic symptoms. The twelve post-traumatic somatic symptoms included in the RPQ are headache, dizziness, nausea, noise sensitivity, fatigue, insomnia, poor concentration, taking longer to think, blurred vision, light sensitivity, double vision, and restlessness (24). For each symptom, a yes/no variable was created. A positive response to a symptom would be counted as YES=1 that it does exist, while a NO=0 indicates that the symptom does not exist. The yes/no responses were summed to provide a simple count of somatic symptoms.

### Sample Processing and DNA Sequencing

Prior to extraction, stool samples were heat inactivated at 65°C-70°C for 1 hour and stored at -80°C. Approximately 250 mg of resulting sample was extracted using the QIAGEN DNeasy PowerSoil Pro Kits (QIAGEN, catalog no. 47016). Sequencing libraries were prepped using the Nextera XT DNA library prep kit and sequenced on a NextSeq 500 sequencer with 2x150 bp paired-end reads. Of the 74 stool samples received, 69 (93.2%) were successfully sequenced. Samples with a percentage of reads identified greater than 0.5%, and complete metadata were used for analysis (n=51).

Shotgun metagenomic reads were trimmed and filtered for host contamination using the KneadData pipeline (https://github.com/biobakery/kneaddata). The resulting metagenomic data was profiled for microbial abundance and metabolic pathways using the MetaPhlAn3(25) and HUMAnN3 databases and tools(26). The resulting relative microbial species abundance was used for downstream analysis in R.

### Microbiome Analysis and Statistics

#### Linear mixed model-based analysis to determine the relative contribution of the microbiome

To determine how much of the variability in post-trauma APNS outcomes is explained by the gut microbiome, we first constructed linear mixed models (LMM) using the lme4 R package(27) for each of the three APNS outcomes of interest (PTSD, Depression Scores and Somatic Symptoms) as a function of relevant clinical covariates (sex, age, body mass index, Race/ethnicity)(28), and also of the arcsine square root-transformed abundance (29) of each microbial species independently. Features identified as significant (FDR corrected p-value <= 0.05) were combined and used to fit a global LMM. The contribution of each feature to the regression line fit by the model was determined by running analysis of variance (ANOVA) and examining the total sum of squares for the fixed effects. The resulting sum of squares ratio for each feature was graphed using WebR (https://github.com/cardiomoon/webr).

#### Mixed-effect random forest analysis of microbiome permutated importance to outcomes at twelve weeks post-trauma

Microbiome data is non-linear and not normally distributed(30, 31). Additionally, such data are characterized by many predictors, which, if combined in the same model (without prefiltering), can prohibit a traditional regression model to converge(32). We have previously demonstrated that tree-based machine learning (ML) approaches such as random forest, which are non-parametric and perform intrinsic feature selection, are very powerful in finding a signal from static and time-resolved microbiome data(33–36). To fully utilize the longitudinal clinical data collected by the AURORA (parent) study for this nested study(1), we assumed, barring perturbation, the gut microbiome is stable over time. Several high-resolution gut microbiome temporal studies have demonstrated that the microbiome is rather stable and displays only small random fluctuations(37–40). We built a mixed-effect random forest (MERF) regression model to predict either PTSD raw score, Depression t-score, or somatic symptoms count at twelve weeks post-trauma. We use either microbial abundance or metabolic pathway abundance and relevant clinical covariates (e.g., BMI, age, sex, and race) as variables in this modeling. The first step of our pipeline split our data into a training and test set. To predict twelve-week post-trauma outcomes, two-week and eight-week post-trauma outcomes were used to train the MERF model. The unseen twelve-week data was used for testing the model. For each APNS outcome, the pipeline was run ten times with ten different random seeds, and model performance, statistics, and outcomes were calculated for each seed. Model performance was evaluated by Root Mean Square Error (RMSE) and correlation of true versus predicted values, which illustrates the model’s accuracy and fit for predicting outcome measurements at twelve weeks post-trauma (Supplementary figure 1). Permuted variable importance calculated and used to evaluate models. Plots summarizing results were generated in R using the ggplot2.

**Figure 1.**
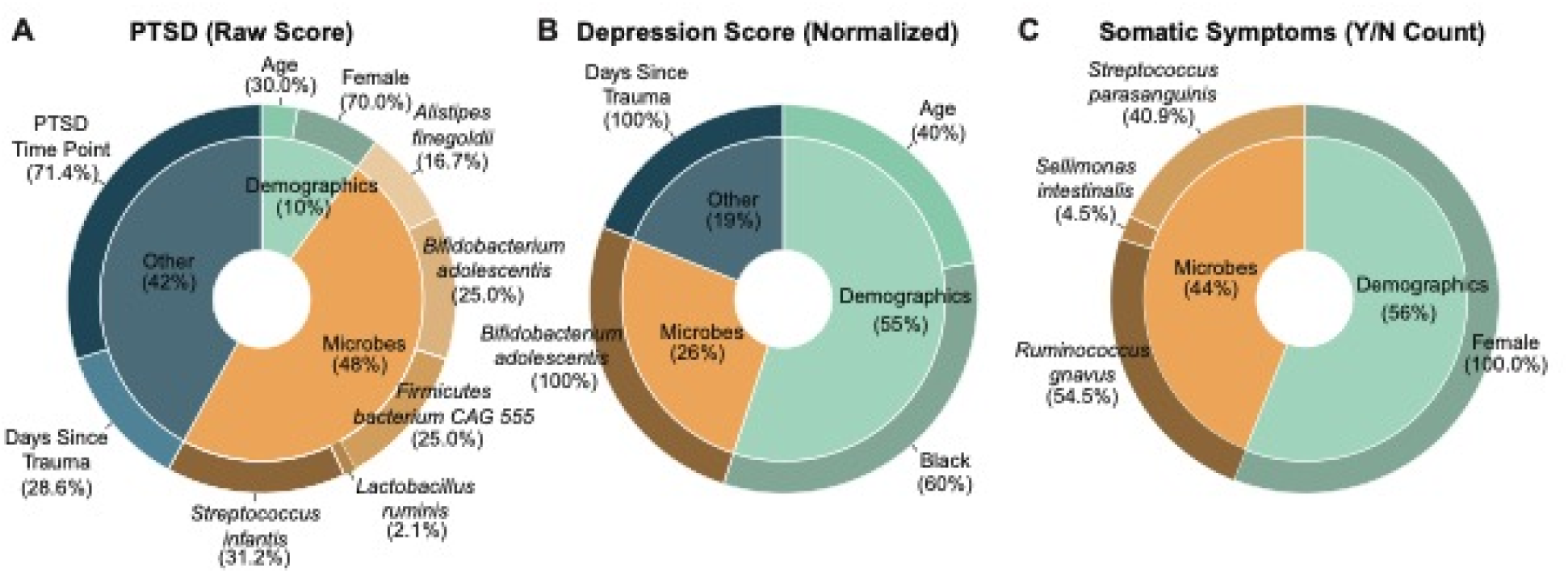
Contribution of microbiome features significantly associated with neuropsychiatric outcomes. After metagenomic sequencing, microbial specie abundance were combined with demographics and select clinical variables (other) in linear mixed effect models for the APNS outcomes at two-, eight-, and twelve weeks post trauma (A) PTSD raw score as determined by the DSM-5 PTSD checklist, (B) depression score as determined by the PROMIS Depression Short Form 8b and (C) somatic symptoms count (Yes/No) based on the Rivermead Post-Concussive Questionnaire. Individual microbial species, demographics, and clinical variables found as significant are displayed in the outer wheels. PTSD time point reflects the time at which the modeled score was taken (either two weeks, eight weeks, or twelve weeks). The length of time between trauma exposure and stool sample collection was included as days since trauma in all models.

## Results

### Characteristics of Study Subjects

Among 2,097 individuals who were approached in the ED regarding this nested study, 106/2,097 (5.1%) agreed to participate, and 74/106 (69.8%) provided a stool sample. Of the stool samples received, 51(68.9%) passed all quality and metadata checks (see methods) and were profiled for microbial abundance and metabolic pathways. From these 51 participants (mean age 52 years; 26 (51%) female) stool samples were received a median of 45 days (range 5 to 182 days) after ED visit for trauma evaluation that conferred entry to the AURORA (parent) study (Table 1). The most common types of trauma were motor vehicle collision (55%, n=28), fall from height (n=9, 18%), and other accidental or targeted/involuntary events (9.8%, n=5). Stool samples originated primarily from White (61%, n=31) and Black (25%, n=13) participants. Of the 31 white participants, 29 (94%) were non-Hispanic White. Microbiome diversity measures show that individuals do not stratify based on standard diversity measures.

**Table 1:** Patient Characteristics by APNS outcome category

|  |  |  | Depression |  |  | PTSD |  |  |
| --- | --- | --- | --- | --- | --- | --- | --- | --- |
| Patient Characteristics |  | Overall, N = 51<br>n (%) | Depression-<br>, N = 35<br>n (%) | Depression+<br>, N = 16<br>n (%) | q-value<br>(adjusted p-value) | PTSD-,<br>N = 31<br>n (%) | PTSD+<br>, N = 20<br>n (%) | q-value<br>(adjusted p-value) |
| Demographics |  |  |  |  |  |  |  |  |
|  | Age in years (at enrollment) | 52 (36, 60) | 52 (36, 59) | 52 (37, 60) | >0.9 | 52 (36, 58) | 50 (34, 61) | >0.9 |
|  | Sex (Female) | 26 (51%) | 19 (54%) | 7 (44%) | >0.9 | 19 (61%) | 7 (35%) | 0.7 |
|  | BMI | 29 (24, 33) | 27 (24, 30) | 33 (29, 40) | 0.11 | 27 (23, 30) | 31 (29, 36) | 0.083 |
|  | White | 31 (61%) | 21 (60%) | 10 (62%) | >0.9 | 18 (58%) | 13 (65%) | >0.9 |
|  | Black | 13 (25%) | 11 (31%) | 2 (12%) | >0.9 | 9 (29%) | 4 (20%) | >0.9 |
|  | Asian | 1 (2.0%) | 1 (2.9%) | 0 (0%) | >0.9 | 1 (3.2%) | 0 (0%) | >0.9 |
|  | other | 6 (12%) | 2 (5.7%) | 4 (25%) | 0.8 | 3 (9.7%) | 3 (15%) | >0.9 |
|  | Hispanic | 7 (14%) | 4 (11%) | 3 (19%) | >0.9 | 4 (13%) | 3 (15%) | >0.9 |
| U.S. Geographic Region |  |  |  |  | >0.9 |  |  | >0.9 |
|  | Midwest Region | 10 (20%) | 5 (14%) | 5 (31%) |  | 5 (16%) | 5 (25%) |  |
|  | Northeast Region | 38 (75%) | 27 (77%) | 11 (69%) |  | 24 (77%) | 14 (70%) |  |
|  | Southern Region | 3 (5.9%) | 3 (8.6%) | 0 (0%) |  | 2 (6.5%) | 1 (5.0%) |  |
| Trauma Event Type (Broad) |  |  |  |  | >0.9 |  |  | >0.9 |
|  | Animal-related | 1 (2.0%) | 1 (2.9%) | 0 (0%) |  | 1 (3.2%) | 0 (0%) |  |
|  | Fall < 10 feet or from unknown height | 9 (18%) | 7 (20%) | 2 (12%) |  | 6 (19%) | 3 (15%) |  |
|  | Fall >= 10 feet | 4 (7.8%) | 2 (5.7%) | 2 (12%) |  | 2 (6.5%) | 2 (10%) |  |
|  | Incident causing traumatic stress exposure to many people | 1 (2.0%) | 1 (2.9%) | 0 (0%) |  | 1 (3.2%) | 0 (0%) |  |
|  | Motor Vehicle Collision | 28 (55%) | 19 (54%) | 9 (56%) |  | 17 (55%) | 11 (55%) |  |
|  | Non-motorized Collision | 3 (5.9%) | 2 (5.7%) | 1 (6.2%) |  | 2 (6.5%) | 1 (5.0%) |  |
|  | other | 5 (9.8%) | 3 (8.6%) | 2 (12%) |  | 2 (6.5%) | 3 (15%) |  |
| Self-Reported Perceived Chance of Dying |  | 6.0 (1.0, 8.5) | 6.0 (1.0, 8.5) | 5.5 (1.8, 8.2) | >0.9 | 6.0 (0.5, 8.5) | 5.5 (1.8, 8.5) | >0.9 |

**Table 2:**
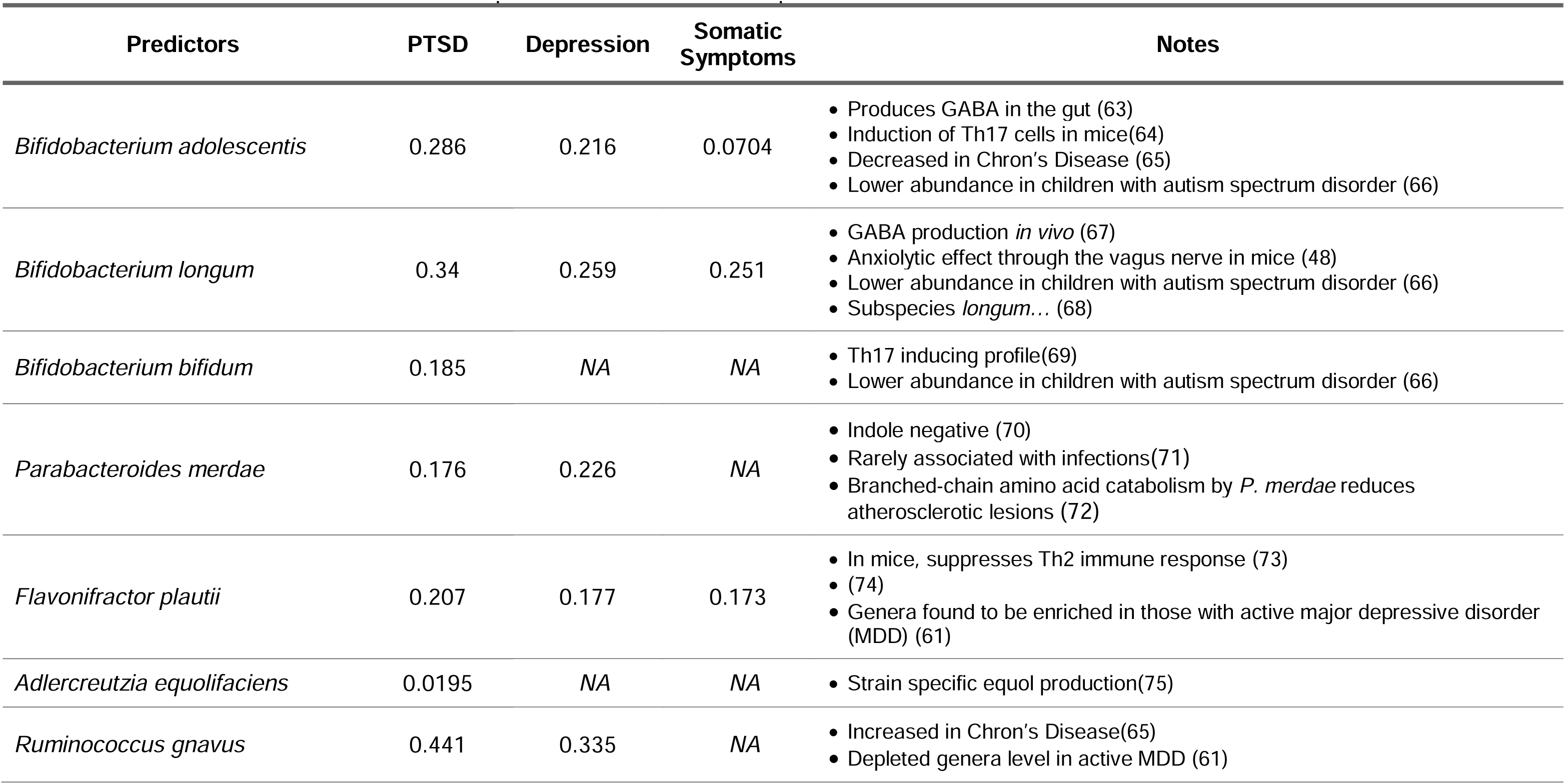
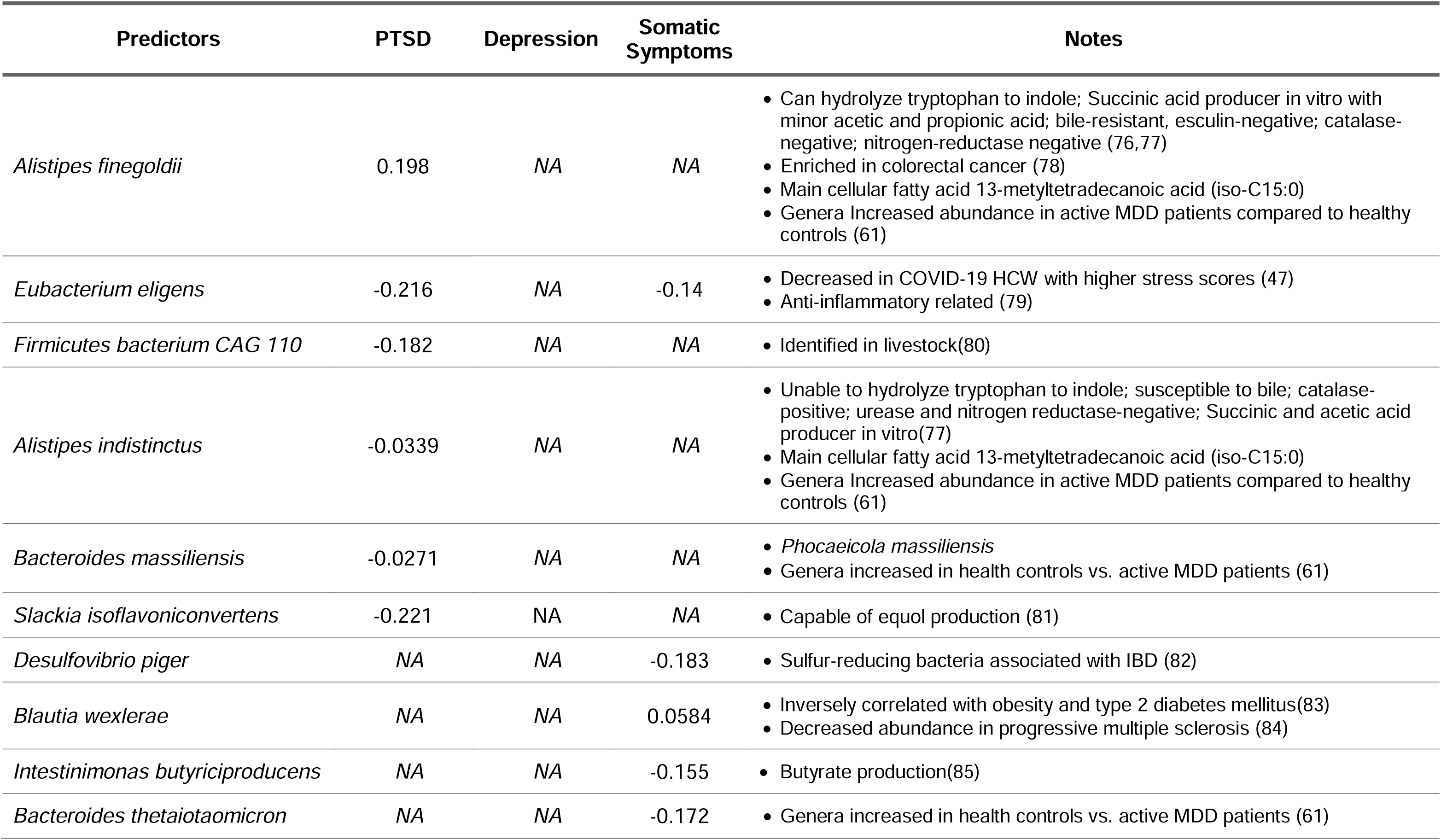
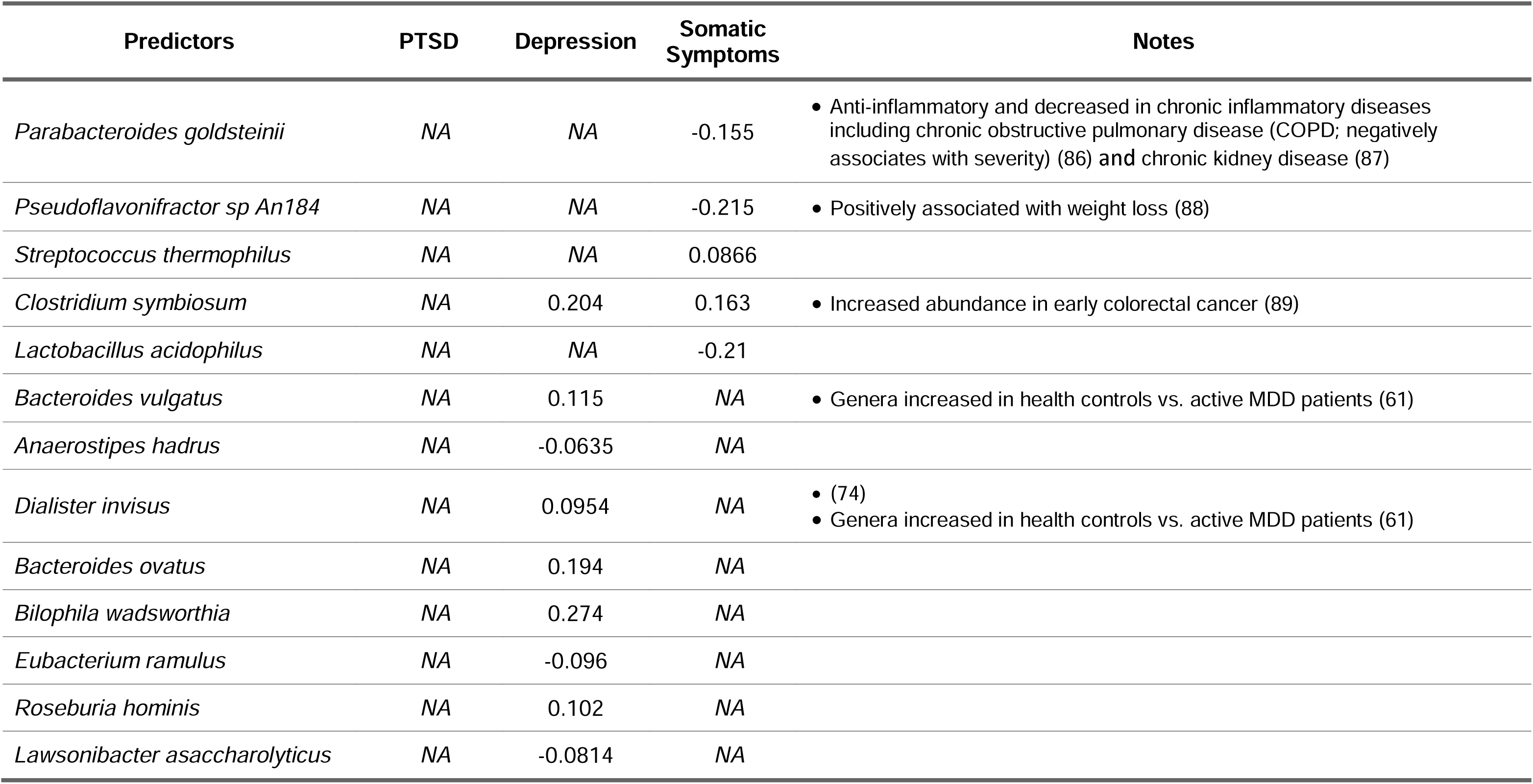
Spearman’s Correlation of Important Microbes with APNS Outcomes

**Table 3:** Spearman’s Correlation of Important Metabolic Pathways with Outcomes

| Predictors | PTSD | Depression | Somatic Symptoms | Notes |
| --- | --- | --- | --- | --- |
| ARG-POLYAMINE-SYN: superpathway of arginine and polyamine biosynthesis | -0.251 | -0.229 | NA | • Enriched patients with CD in remission (90) |
| CITRULBIO-PWY: L-citrulline biosynthesis | 0.346 | 0.298 | 0.261 | • Enriched patients with CD in remission (90) |
| CALVIN-PWY: Calvin Benson Bassham cycle | -0.269 | -0.315 | NA | • Enriched patients with CD in remission (90) |
| ARGININE-SYN4-PWY: L-ornithine de novo biosynthesis | 0.201 | 0.165 | 0.139 |  |
| CENTFERM-PWY: pyruvate fermentation to butanoate | -0.322 | -0.26 | -0.168 |  |
| BIOTIN-BIOSYNTHESIS-PWY: biotin biosynthesis I | 0.181 | 0.127 | NA |  |
| BRANCHED-CHAIN AA-SYN-PWY: superpathway of branched amino acid biosynthesis | -0.127 | -0.00818 | NA |  |
| ANAEROFRUCAT-PWY: homolactic fermentation | -0.0199 | NA | NA |  |
| ASPASN-PWY: superpathway of L-aspartate and L-asparagine biosynthesis | -0.0404 | 0.0571 | 0.037 |  |
| COA-PWY-1: coenzyme A biosynthesis II | -0.0454 | NA | -0.053 |  |
| ARGSYNBSUB-PWY: L-arginine biosynthesis II (acetyl cycle) | -0.0768 | -0.131 | NA |  |
| ARGSYN-PWY: L-arginine biosynthesis I (via L-ornithine) | -0.0579 | -0.105 | NA |  |
| 1CMET2-PWY: N<sup>10</sup> formyl tetrahydrofolate biosynthesis | NA | -0.0885 | NA | • Depleted in UC in remission (90) |
| ANAGLYCOLYSIS-PWY: glycolysis III (from glucose) | NA | -0.048 | NA |  |
| ARGORNPROST-PWY: arginine, ornithine and proline interconversion | NA | 0.0264 | NA |  |

### Gut Microbiome Features associated with APNS

Linear mixed effect models and ANOVA adjusting for sociodemographic characteristics were used to assess for associations between microbiome characteristics and APNS symptom severity (As noted above, analyses were accounted for sociodemographic factors because of the known association between socioeconomic status and diet (36)). Gut microbiome characteristics accounted for 48%, 26%, and 44% of the variation in PTSD, depressive, and somatic symptoms after trauma, respectively (Figure 1). The abundance of *Firmicutes bacterium CAG:555, Bifidobacterium adolescentis,* and the pro-inflammatory *Streptococcus infantis* were associated with PTSD symptom severity. The abundance of *B. adolescentis* was associated with depressive symptom severity (Figure 1B). *Ruminococcus gnavus* and *Streptococcus parasanguinus* were associated with somatic symptom severity (Figure 1C).

### Post-Trauma Neuropsychiatric Outcomes are Predicted by Microbial Abundance

While the above LMM approach is useful to assess total variance accounted for by microbiome characteristics and is common in the field, it has several limitations, including the inability to evaluate microbial abundances in the context of the entire microbiome and the limited ability to account for inter-individual microbiome differences(32). Therefore, we used a complementary machine learning approach to train and test models examining associations between gut microbiome characteristics and posttraumatic outcomes. We have previously used this approach and have determined its optimality for inferring biologically relevant host-microbe interactions from cross-sectional and longitudinal microbiome data (see methods sections for further justification)(33–36,41–43).

A Mixed-Effect Random Forest (MERF) regression model was used to identify microbiome features associated with PTSD, depression, and somatic symptom burden at twelve weeks post-trauma. Our findings using this approach confirmed and expanded on findings from the linear mixed-effect models. Bacterial species identified in linear mixed effects modeling, specific species of the *Alistipes, Bifidobacterium*, and *Ruminococcus* genera were also found as important predictors by the MERF model (Figure 2). The models also identified more species as contributors to each outcome, with many similar species in the top 15 predictors identified by the three models. *B. adolescentis*, *B. longum*, and *Flavonifractor plautii* were among the top five predictors for all three APNS outcomes (Figure 2). Across our models, both *Bifidobacterium* species and *F. plautii* showed similar trends of increased abundance being informative of higher scores (Supplemental figure 3). *Ruminococcus gnavus* was found to be among the top 15 predictors only for PTSD (ranked 7^th^) and depression (ranked 14^th^), with increases in abundance correlating with disease outcomes (Figure 2). Decreases in certain species were noted to cluster by individual outcomes for the most part (Figure 2D).

**Figure 2.**
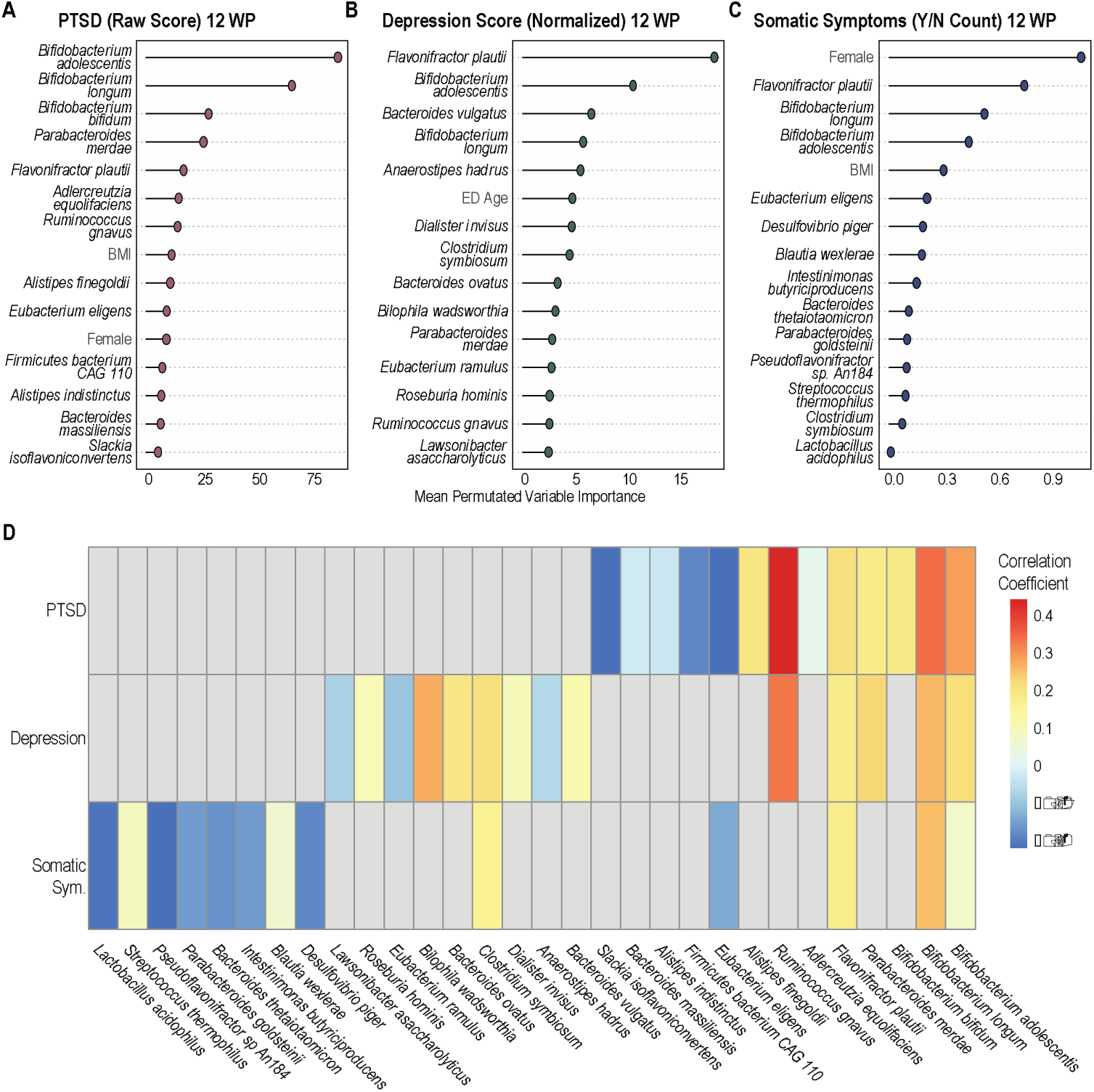
Mixed-Effect Random Forest (MERF) Regression Models Using Microbial Abundance and Clinical Covariates to Predict Neuropsychiatric Outcomes. MERF models combining microbial abundance data with clinical and demographic features demonstrate the importance of microbial features in predicting outcomes. Permutated importance analysis of model outcomes shows the top 15 features contributing to predictions of (A) PTSD raw score, (B) depression normalized score, and (C) somatic symptoms count (yes/no) are mostly microbial species. (D) Heat map showing correlation coefficients of the top 30 contributing species show common species that contribute to all 3 outcomes (increased abundance) and species which associate with individual outcomes (mostly decreased abundance).

### Metabolic Pathway Profiling Identifies Novel Gut-Brain-Axis Interface with APNS Outcomes

Species abundance, although informative of outcomes, does not directly provide context for the functional roles of associated microbes. By analyzing metabolic pathway abundances encoded in the metagenomic data, we can gain insight into the microbial products, metabolites, and functions that may associate with each outcome of interest, thus shedding light on possible mechanistic links. To this end, we employed the same MERF-based pipeline to examine associations between the abundance of microbially-encoded metabolic pathways and PTSD, depressive, and somatic symptom severity twelve weeks after trauma. The Calvin-Benson- Bassham cycle, a common CO2 fixation pathway in autotrophic bacteria, was identified as one of the top three predictors of PTSD and depressive symptom severity (i.e., cycle was reduced in samples with higher scores (Figures 3A and 3B)). Amino acid biosynthesis pathways were a leading predictor for all three APNS outcomes, with the L-citrulline biosynthesis pathway identified as one of the top two predictors for all APNS outcomes. The super pathway of arginine and polyamine biosynthesis was identified as the top predictor for the PTSD model and as the third most important pathway for the depression model, correlating negatively with PTSD and depression scores. Furthermore, all three models identified pathways involving arginine, ornithine, and citrulline, three amino acids that are often interconverted (Figure 3). *De novo* biosynthesis of L-ornithine, which can be interconverted from ornithine and urea to arginine and water, was identified by all models and appeared increased in patients with higher scores. Increased abundance of L-citrulline biosynthesis was also associated with higher scores of PTSD, depression, and somatic symptoms count.

**Figure 3.**
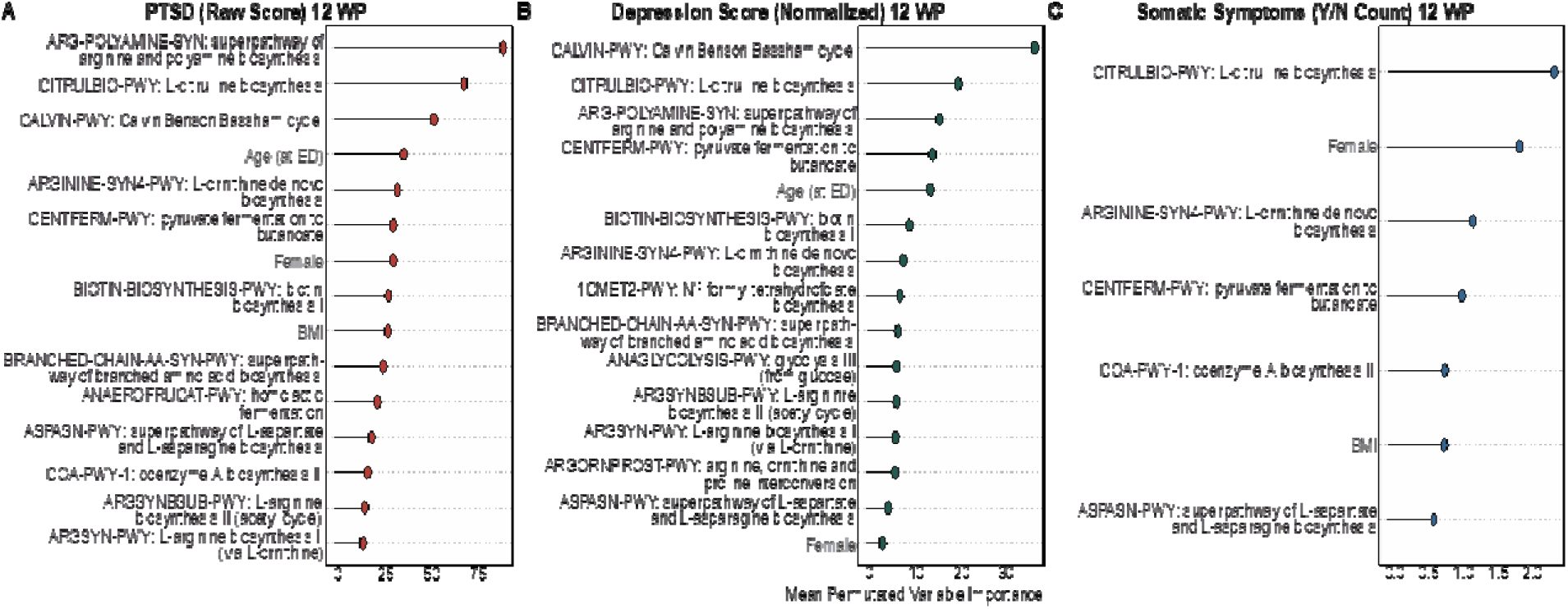
Mixed-Effect Random Forest (MERF) Regression Models Using Microbial Metabolic Pathways and Clinical Covariates to Predict Neuropsychiatric Outcomes. MERF models combining microbial metabolic pathway abundance data with clinical and demographic features demonstrate the importance of microbial pathways in predicting outcomes for (A) PTSD raw score, (B) depression normalized score, and (C) somatic symptoms count (yes/no). The top 15 features from analysis of permutated importance on model outcomes are displayed for PTSD and depression. Our model found only 7 significant features for somatic symptoms.

**Figure 4.**
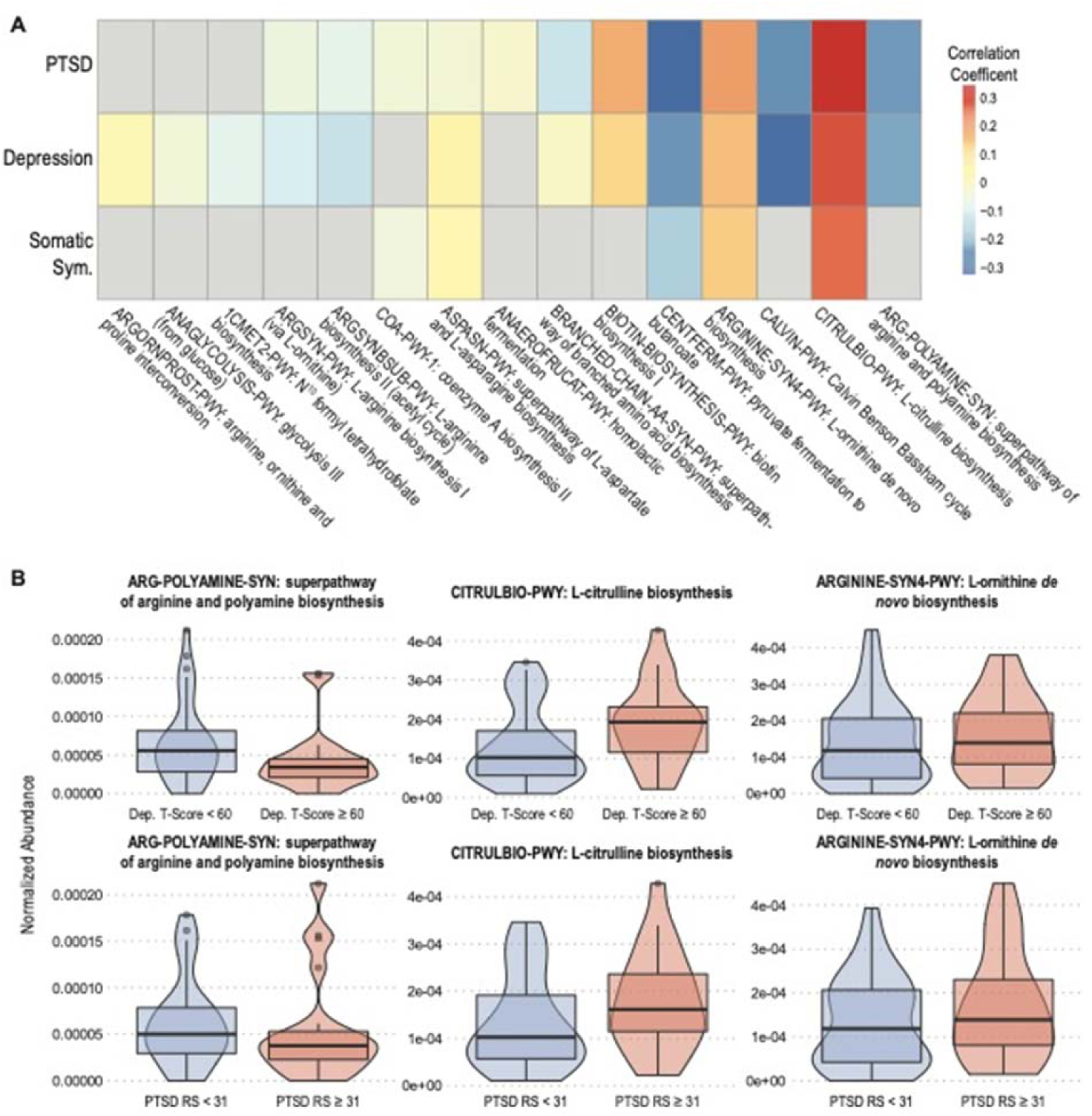
Important metabolic pathways for PTSD and depression involve arginine, citrulline and ornithine. Heat map (A) of correlation coefficients for the top 15 metabolic pathways contributing to PTSD, depression, and somatic symptoms from MERF analysis show significant contributions by amino acid and polyamine biosynthesis pathways. (B) Violin plots showing differences in contributions of arginine, citrulline, and ornithine biosynthesis pathways for depression (top graphs) and PTSD (bottom graphs). Blue violins indicate no diagnosis (PTSD raw score ≤31; Depression T-Score < 60 indicating none to mild depression) red indicates PTSD or depression diagnosis (PTSD RS > 31; Depression T-Score ≥60 indicating moderate to severe depression).

**Figure 5.**
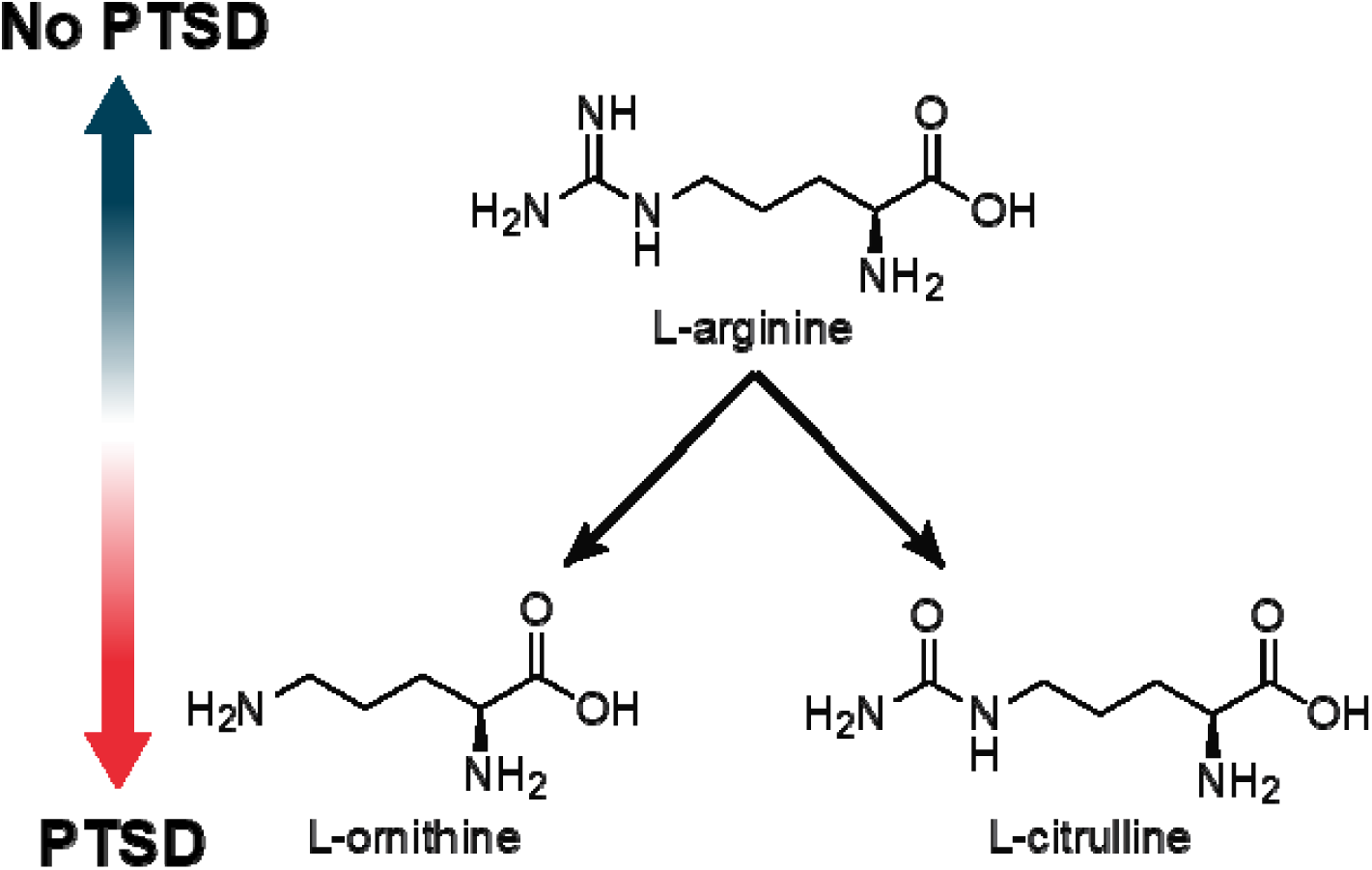
Arginine is converted into citrulline and ornithine commonly in the host and the microbiome. The ratio of arginine to citrulline and ornithine in PTSD patients has previously been found to negatively correlated with PTSD. We find that this negative correlation in the metabolic pathways encoded for in the microbiome of trauma survivors.

Given the strength of findings around pathways affecting global arginine levels, we next sought to identify microbes responsible for changes in these pathways. Focusing on ornithine, citrulline, and arginine, we examined the contribution of individual genera to each pathway (Tables 4-6) to PTSD and depression. Somatic symptoms were not further analyzed as the simple count nature of the variable was not appropriate for this analysis. We observed that in individuals with PTSD or depression, *Escherichia* had an increased contribution to all pathways involving arginine and ornithine biosynthesis. However, the contribution of *E. coli* to L-citrulline biosynthesis was decreased in those with PTSD and increased in those with depression. In patients with PTSD or depression, the *Ruminococcus* genus had a decreased contribution to L-arginine biosynthesis via both L-ornithine and the acetyl cycle. *Alistipes*, *Flavonifactor* and *Faecalibacterium* genera contributed more to the L-arginine biosynthesis via ornithine in individuals with diagnosed PTSD or depression.

**Table 4:**
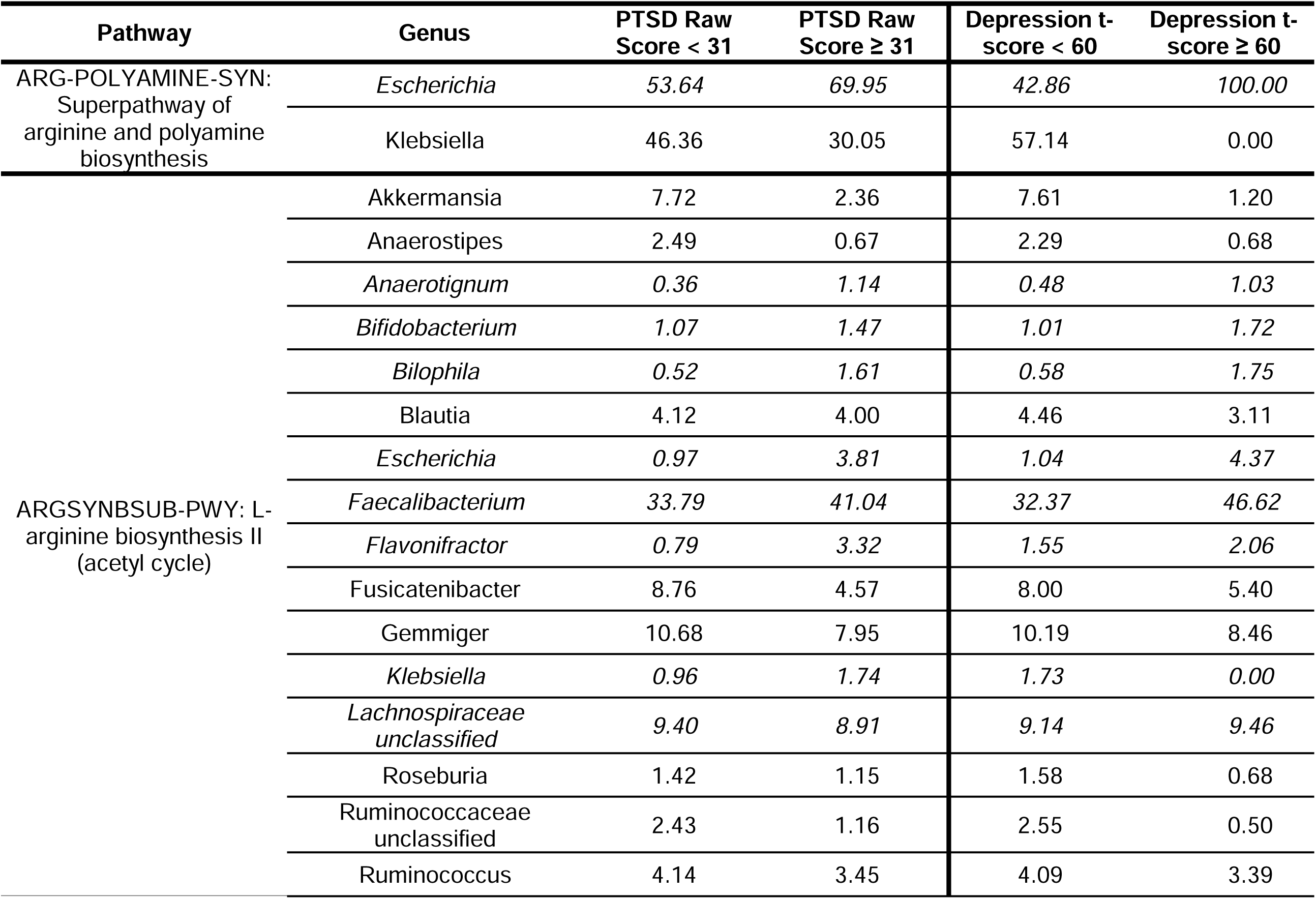
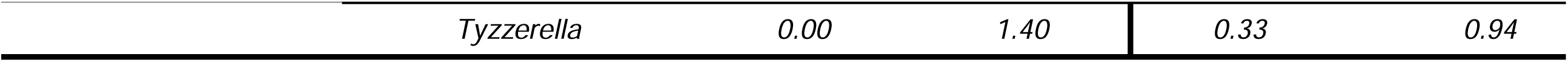
Microbial genus contribution to arginine-related metabolic pathways stratified by PTSD and depression diagnoses

**Table 5:** Microbial genus contribution to ornithine-related metabolic pathways stratified by PTSD and depression diagnoses

| Pathway | Genus | PTSD Raw Score < 31 | PTSD Raw Score ≥ 31 | Depression t-score < 60 | Depression t-score ≥ 60 |
| --- | --- | --- | --- | --- | --- |
| ARGININE-SYN4-PWY: L-ornithine *de novo* biosynthesis | *Bacteroides* | 38.78 | 29.26 | 37.40 | 28.63 |
|  | *Catenibacterium* | 13.22 | 0.14 | 7.15 | 4.90 |
|  | *Escherichia* | 12.28 | 25.78 | 12.20 | 29.42 |
|  | *Klebsiella* | 5.21 | 4.50 | 8.41 | 0.00 |
|  | *Parabacteroides* | 28.82 | 35.58 | 33.49 | 31.06 |
|  | *Paraprevotella* | 1.68 | 4.75 | 1.35 | 5.99 |
| ARGSYN-PWY: L-arginine biosynthesis I (via L-ornithine) | *Alistipes* | 3.21 | 16.69 | 3.92 | 18.18 |
|  | *Anaerostipes* | 2.60 | 0.38 | 2.40 | 0.31 |
|  | *Bifidobacterium* | 1.28 | 1.47 | 1.21 | 1.65 |
|  | *Blautia* | 3.95 | 3.29 | 4.29 | 2.46 |
|  | *Escherichia* | 1.36 | 3.40 | 1.45 | 3.65 |
|  | *Faecalibacterium* | 38.95 | 39.19 | 37.33 | 42.50 |
|  | *Flavonifractor* | 0.72 | 2.79 | 1.56 | 1.56 |
|  | *Fusicatenibacter* | 9.18 | 3.56 | 8.30 | 4.12 |
|  | *Gemmiger* | 11.38 | 7.32 | 10.89 | 7.41 |
|  | *Klebsiella* | 0.55 | 0.77 | 0.96 | 0.00 |
|  | *Lachnospiraceae unclassified* | 9.42 | 7.48 | 9.13 | 7.64 |
|  | *Roseburia* | 2.06 | 1.20 | 2.30 | 0.55 |
|  | *Ruminococcaceae unclassified* | 2.32 | 1.02 | 2.49 | 0.41 |
|  | *Ruminococcus* | 4.51 | 3.05 | 4.43 | 2.89 |

**Table 6:**
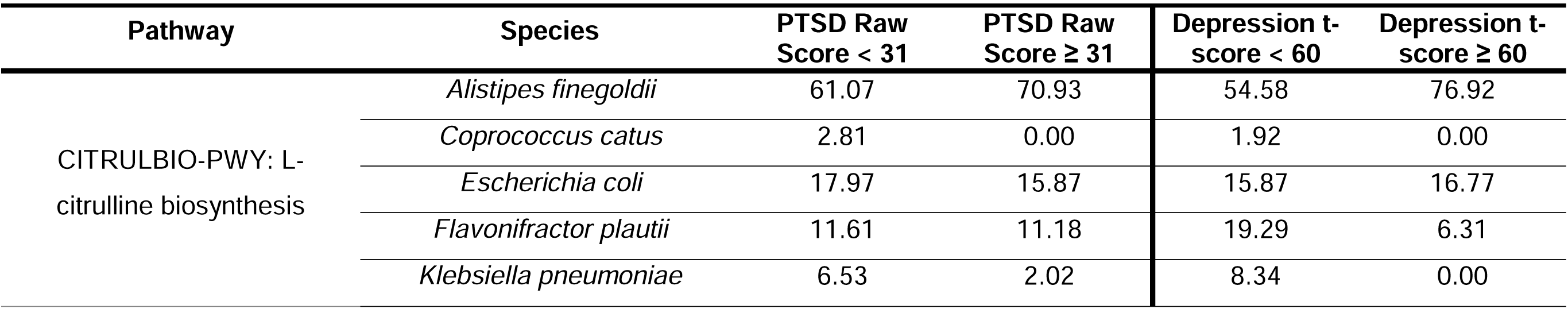
Microbial genus contribution to citrulline-related metabolic pathways stratified by PTSD and depression diagnoses

| Pathway | Species | PTSD Raw Score < 31 | PTSD Raw Score ≥ 31 | Depression t-score < 60 | Depression t-score ≥ 60 |
| --- | --- | --- | --- | --- | --- |
| CITRULBIO-PWY: L-citrulline biosynthesis | *Alistipes finegoldii* | 61.07 | 70.93 | 54.58 | 76.92 |
|  | *Coprococcus catus* | 2.81 | 0.00 | 1.92 | 0.00 |
|  | *Escherichia coli* | 17.97 | 15.87 | 15.87 | 16.77 |
|  | *Flavonifractor plautii* | 11.61 | 11.18 | 19.29 | 6.31 |
|  | *Klebsiella pneumoniae* | 6.53 | 2.02 | 8.34 | 0.00 |

## Discussion

We have demonstrated here that gut microbiome characteristics of trauma survivors were associated with the development of APNS, in this pilot longitudinal study of individuals enrolled in the ED after trauma exposure. Our analysis first suggested that overall inter-individual differences in gut microbiome taxonomy accounts for a substantial fraction (20-48%) of the differences in APNS outcomes. This finding is similar to what is attributable to known APNS outcome-associated clinical and demographic covariates (e.g., sex and race/ethnicity)(44). We leveraged ML modeling to identify specific bacterial species and encoded metabolic pathways with previously established pro- and anti-inflammatory properties that are predictive of APNS development. Our analysis highlights the most prevalent APNS outcome-associated gut microbiome-encoded pathways are those leading to the biosynthesis of arginine, ornithine, and citrulline. These pathways can possibly affect global arginine levels in the body, a biomarker that has been previously associated with PTSD(45). To our knowledge, ours is the first study to longitudinally evaluate the link between the gut microbiome and APNS outcomes while providing mechanistic links along the microbiome-gut-brain axis.

Our initial analyses by simple linear mixed-effect models demonstrated the significant contribution of microbiota to APNS outcomes in comparison to clinical covariates and demographic characteristics. The proportion of the gut microbiome accounting for variability in each APNS outcome (26-48%) is of the same order of magnitude as demographic characteristics (10-56%) that are already known to associate with each outcome(44). However, this analysis required transformation of microbiome abundance variables and the use of a two-tiered approach to select outcome-associated features, which is suboptimal. Although common, this approach neither accounts for the inter-personal variation of the microbiome nor evaluates the effect of microbial species together(46).

While there is evidence for dramatic changes in the gut microbiome after trauma within 72 hours(16), almost no research describing long-term changes in gut microbiota after acute trauma currently exists. One study on PTSD in frontline healthcare workers suggests that long-term gut microbiome dysbiosis induced by stress is sustained for months and predisposes individuals to recurring PTSD(47). Thus, we built our pipeline assuming the microbiome at our sampling point is representative of the patient’s microbiome at the longitudinal post-trauma outcome measurement points, which we hypothesize associates with target outcomes of developing APNS in prolonged periods after trauma.

ML models such as Mixed-Effect Random Forest (MERF) models are capable of accounting for interpersonal variation, evaluating microbial abundances in the context of the entire microbiome, and, importantly, because no scaling is required, it can be used with different data types (e.g., categorical, numerical, proportion, etc.)(32)Our MERF models identified more species as significant contributors to our outcomes than the linear models and microbes previously associated with neuropsychiatric disorders, specifically *Bifidobacterium*, *Alistipes*, and *Flavonifractor* species. Further, the model identified *F. plauti*, *E. eligens*, *E. ramulus*, *R. homins*, *C. sybiosum,* and *Blautia* species which are members of the *Clostridium* Cluster IV and XIVa, two clusters of known beneficial bacteria for inflammatory bowel disease and gut health(56–58). Several of these are short-chain fatty acid producers and have been associated with reduced inflammation in humans (59, 60). Additionally, they are known to promote several anti-inflammatory immune signatures, such as regulatory T-cell expansion *in vivo* (61, 62).

Our models based on species abundance identified *Bifidobacterium* species and *Flavonifractor plautii* as the most important factors for predicting all three APNS outcomes. Specifically, we found higher abundances of *B. adolescentis, B. longum,* and *B. bifidum* were associated with higher PTSD and depression scores, which contradicts some literature regarding positive relationships between *Bifidobacterium spp* and neuropsychiatric outcomes(48–50). Contrary to this, many other microbiome-based studies of neuropsychiatric disorders, including major depressive disorder (MDD), schizophrenia, and anxiety, have failed to find similar associations(51–53). Indeed, for MDD, various investigators have found both increased and decreased abundances of *Bifidobacterium spp* to be associated with clinical disease(53–55). Our analysis identifying increased abundances of *Bifidobacterium spp* as being associated with APNS outcomes adds more contradiction to the literature but highlights the importance of looking beyond species abundances in microbiome studies.

Alone, species-based associative studies can be confounding and limit clearer investigations into biological mechanisms. We have previously shown that also analyzing microbial metabolic pathways can reveal valuable insight into possible biologic impacts of the gut microbiome on clinical outcomes(34, 41). As with our previous work, our combined analysis approach here, leveraging metagenomic sequencing and ML-based pipelines on both species’ abundance and metabolic pathways, identified a unique microbially-based mechanism with the biosynthesis of arginine/citrulline/ornithine as highly informative of the development of all three APNS outcomes. This is the first described biologic link in APNS attributable directly to the microbiome-gut-brain axis and microbially produced metabolites.

PTSD patients have been shown to have decreased levels of arginine and increased levels of ornithine and citrulline in peripheral blood(45). The ratio of arginine to its two main catabolic products, ornithine, and citrulline, is used as a readout of nitric oxide (NO) capacity and is referred to as the global arginine bioavailability ratio (GABR)(56). Bersani et. al. sought to examine NO production in PTSD patients via the GABR as arginine is the sole nitrogen source of NO synthesis. They found that GABR was negatively correlated with PTSD, which they reasoned was indicative of dysfunctional NO synthase. The directionality observed by Bersani and authors mirrors the directionality of arginine-related metabolic pathways in our analysis.

Altered arginine metabolism has also been implicated in other neuropsychiatric disorders, such as schizophrenia and MDD(57, 58). The influence of altered arginine metabolism is likely multidimensional and spans multiple mechanisms of action, including through arginine vasopressin or NO(59, 60). Identifying microbial sources of these potential mechanisms in specific neuropsychiatric conditions is an important first step towards enabling research on interventions or therapies.

In our species trained MERF pipeline, *Flavonifactor plautii,* a flavonoid-degrading bacterium often found in the human gut microbiome, was the only species among the top 5 contributors to all three APNS outcomes that affect GABR. This genus contributes to the L-arginine biosynthesis pathway via ornithine in those with PTSD and via acetyl in those with PTSD or depression. From our metabolic pathway models, the contribution of *Ruminococcus*, *Alistipes* and *Flavonifactor* genera to L-arginine biosynthesis via ornithine was identified as increased in individuals with PTSD or depression. Although the contribution of these three species were not ranked as highly as *Bifidobacterium spp* in our species trained MERF model, by analyzing microbial metabolic pathways, we see that their roles in PTSD and depression may be far more significant.

Our findings also provide an added layer of contextual insight into seemingly contradictory findings from prior research on this topic. In addition to being increased in Crohn’s disease, the *Ruminococcus* genera have been identified as decreased in those with MDD in multiple studies(50, 61). Although we find *R. gnavus* abundance increases with depression score, the contribution of the *Ruminococcus* genera to arginine biosynthesis through the acetyl cycle (ARGSYNBSUB-PWY) and via ornithine (ARGSYN-PWY) was lower in those with depression compared to those without, implying species specific effects may also be at work.

Likewise, *Alistipes* have been found to be both increased(61) and decreased in patients with MDD(50, 62) as compared to healthy controls, suggesting genus-based examination of *Alistipes* may not be sufficient. *A. finegoldii* and *A. indistinctus*, were both *identified* by our microbial model as informative of PTSD score yet with opposite directions. Furthermore, *A. finegoldii* abundance was positively associated with PTSD scores and specifically had a higher contribution to L-citrulline biosynthesis in those with PTSD or depression. We also found that the Alistipes genera contributed more to ornithine-related pathways in those with PTSD and depression. Thus, by combining analyses of species abundance with microbial metabolic pathways, we can better dissect the functional contributions of the microbiota.

The emerging role of polyamines in neuropsychiatric disorders opens a door to a better understanding of the complex pathophysiology of these disorders. The ability of microbiota to produce polyamines and known associations between polyamine-producing microbes and neuropsychiatric disorders highlight the importance of the microbiome-gut-brain axis in human health. Our novel finding of the gut microbiome contributing to alterations in GABR pathways in our studied outcomes is the first direct mechanistic link between the gut microbiome and APNS. This result may provide some indirect evidence of a biological link for APNS along the microbiome-gut-brain axis via microbially generated metabolites.

### Strengths and Limitations

We are the first to delve into the predictiveness of the microbiome to the core components of APNS. Although the sample size of our study was limited, our employment of ML methods and metagenomic profiling enabled us to maximize the utility and richness of data from the samples we had. Furthermore, the AURORA (parent) study ensured a comprehensive, thorough, and standardized system of scoring APNS outcomes. We acknowledge that single time point collections are less ideal for investigating mechanisms driving potentially prolonged and dynamic outcomes. Additionally, we encountered logistical difficulties with recruitment and sample collection due to COVID-19 restrictions. Future work will aim to expand on our preliminary findings with a larger cohort and more sampling time points, including sampling closer to time of trauma. Lastly, although we approached these APNS diagnoses as discrete outcomes, it is well known that there is much overlap between them. Traditional APNS classification evolved from the realms of specific medical specialties and thus are not indexed to specific biological processes or basis(1). This biologic overlap may be responsible for the overlap in some features identified by our modeling. A study with a larger sample size may be able to tease apart the overlap of these outcomes.

## Conclusions

APNS can have devastating long-term consequences for patients who have already suffered trauma, but APNS may be preventable. Our nested study, using a subset of the AURORA cohort, demonstrated the importance of the microbiome in influencing APNS development. While more work is needed, we are the first to describe a possible biologic link between the gut microbiome and post-trauma outcomes through arginine metabolism and global arginine pathways, which have already been associated with PTSD and other neuropsychiatric disorders. This discovery opens avenues for investigating prevention and treatment strategies through both targeted therapies and microbiome-based interventions. Our findings provide some evidence of a biological link for APNS along the microbiome-gut-brain axis via microbially generated metabolites.

## Data and code availability

Microbiome sequencing data is deposited in the Short Read Archive (SRA) under accession no.XXXXXX. Processed data and code to reproduce the results are available at XXXXXX. All data produced in the present study are also available upon reasonable request to the authors.

## Data Availability

All data produced in the present study are available upon reasonable request to the authors

## Acknowledgments

We would like to thank the trauma survivors who participated in the AURORA study and in our microbiome sub-study. Their time and effort during a challenging period of their lives make our efforts to improve recovery for future trauma survivors possible. This work was supported by R01 AG067483-01 to J.H. and by the CDMRP PRMP W81XWH2020013 to V.B. We would also like to thank Dr. Cindy Lai for her assistance with preparing and editing this manuscript.

The AURORA (parent) study was supported by NIMH under U01MH110925, the US Army MRMC, One Mind, and The Mayday Fund. The content is solely responsibility of the authors and does not necessarily represent the official views of any of the funders. Data and/or research tools used in the preparation of this manuscript were obtained from the National Institute of Mental Health (NIMH) Data Archive (NDA). NDA is a collaborative informatics system created by the National Institutes of Health to provide a national resource to support and accelerate research in mental health. Dataset identifier(s): NIMH Data Archive Digital Object Identifier (DOI) 10.15154/1528593. This manuscript reflects the views of the authors and may not reflect the opinions or views of the NIH or of the Submitters submitting original data to NDA.

## Disclosures

Dr. Neylan has received research support from NIH, VA, and Rainwater Charitable Foundation, and consulting income from Boehringer Ingelheim and Jazz Pharmaceuticals.

In the last three years Dr. Clifford has received research funding from the NSF, NIH and LifeBell AI, and unrestricted donations from AliveCor Inc, Amazon Research, the Center for Discovery, the Gates Foundation, Google, the Gordon and Betty Moore Foundation, MathWorks, Microsoft Research, Nextsense Inc, One Mind Foundation, the Rett Research Foundation, and Samsung Research. Dr Clifford has financial interest in AliveCor Inc and Nextsense Inc. He also is the CTO of MindChild Medical and CSO of LifeBell AI and has ownership in both companies. These relationships are unconnected to the current work.

Dr. Rauch reports grants from NIH during the conduct of the study; personal fees from SOBP (Society of Biological Psychiatry) paid role as secretary, other from Oxford University Press royalties, other from APP (American Psychiatric Publishing Inc.) royalties, other from VA (Veterans Administration) per diem for oversight committee, and other from Community Psychiatry/Mindpath Health paid board service, including equity outside the submitted work; other from National Association of Behavioral Healthcare for paid Board service; other from Springer Publishing royalties; and Leadership roles on Board or Council for SOBP, ADAA (Anxiety and Depression Association of America), and NNDC (National Network of Depression Centers).

Dr. Sheikh has received funding from the Florida Medical Malpractice Joint Underwriter’s Association Dr. Alvin E. Smith Safety of Healthcare Services Grant; Allergan Foundation; the NIH/NIA-funded Jacksonville Aging Studies Center (JAX-ASCENT; R33AG05654); and the Substance Abuse and Mental Health Services Administration (1H79TI083101-01); and the Florida Blue Foundation.

Dr. Jones has no competing interests related to this work, though he has been an investigator on studies funded by AstraZeneca, Vapotherm, Abbott, and Ophirex.

Dr. McLean served as a consultant for Walter Reed Army Institute for Research and for Arbor Medical Innovations.

In the past 3 years, Dr. Kessler was a consultant for Cambridge Health Alliance, Canandaigua VA Medical Center, Holmusk, Partners Healthcare, Inc., RallyPoint Networks, Inc., and Sage Therapeutics. He has stock options in Cerebral Inc., Mirah, PYM, and Roga Sciences.

Dr. Koenen’s research has been supported by the Robert Wood Johnson Foundation, the Kaiser Family Foundation, the Harvard Center on the Developing Child, Stanley Center for Psychiatric Research at the Broad Institute of MIT and Harvard, the National Institutes of Health, One Mind, the Anonymous Foundation, and Cohen Veterans Bioscience. She has been a paid consultant for Baker Hostetler, Discovery Vitality, and the Department of Justice. She has been a paid external reviewer for the Chan Zuckerberg Foundation, the University of Cape Town, and Capita Ireland. She has had paid speaking engagements in the last three years with the American Psychological Association, European Central Bank. Sigmund Freud University – Milan, Cambridge Health Alliance, and Coverys. She receives royalties from Guilford Press and Oxford University Press.

Dr. Bucci reports funding from the National Institute of Health, the Department of Defense, and the Bill & Melinda Gates Foundation for unrelated work. Dr. Bucci is also supported by a Sponsored Research Agreement from Vedanta Biosciences, Inc, which is unrelated to the present work.

## Supplemental Figures

**Supplemental Figure 1.**
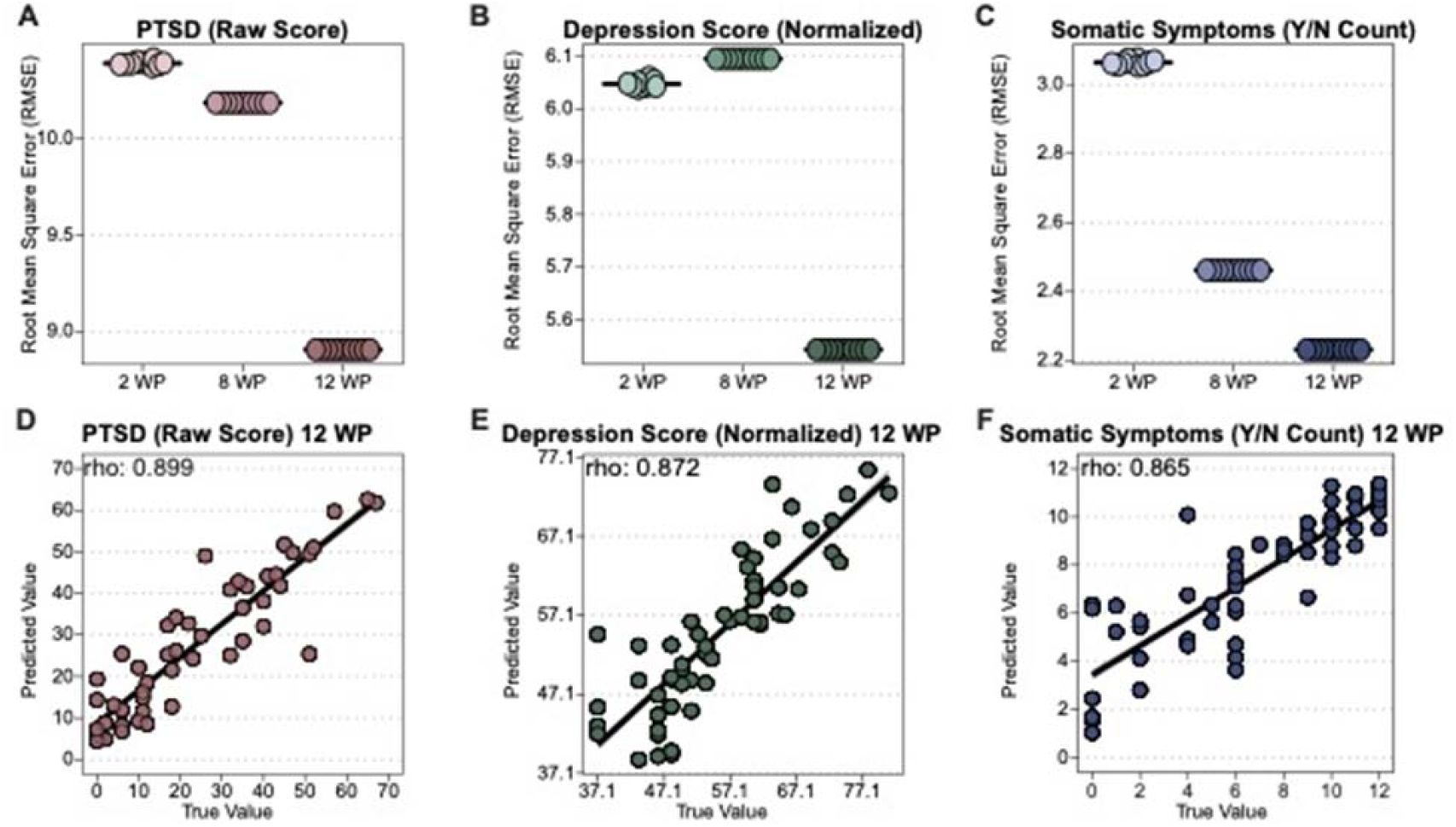
RMSE and correlation graphs for MERF models trained with microbial abundance data.

**Supplemental Figure 2.**
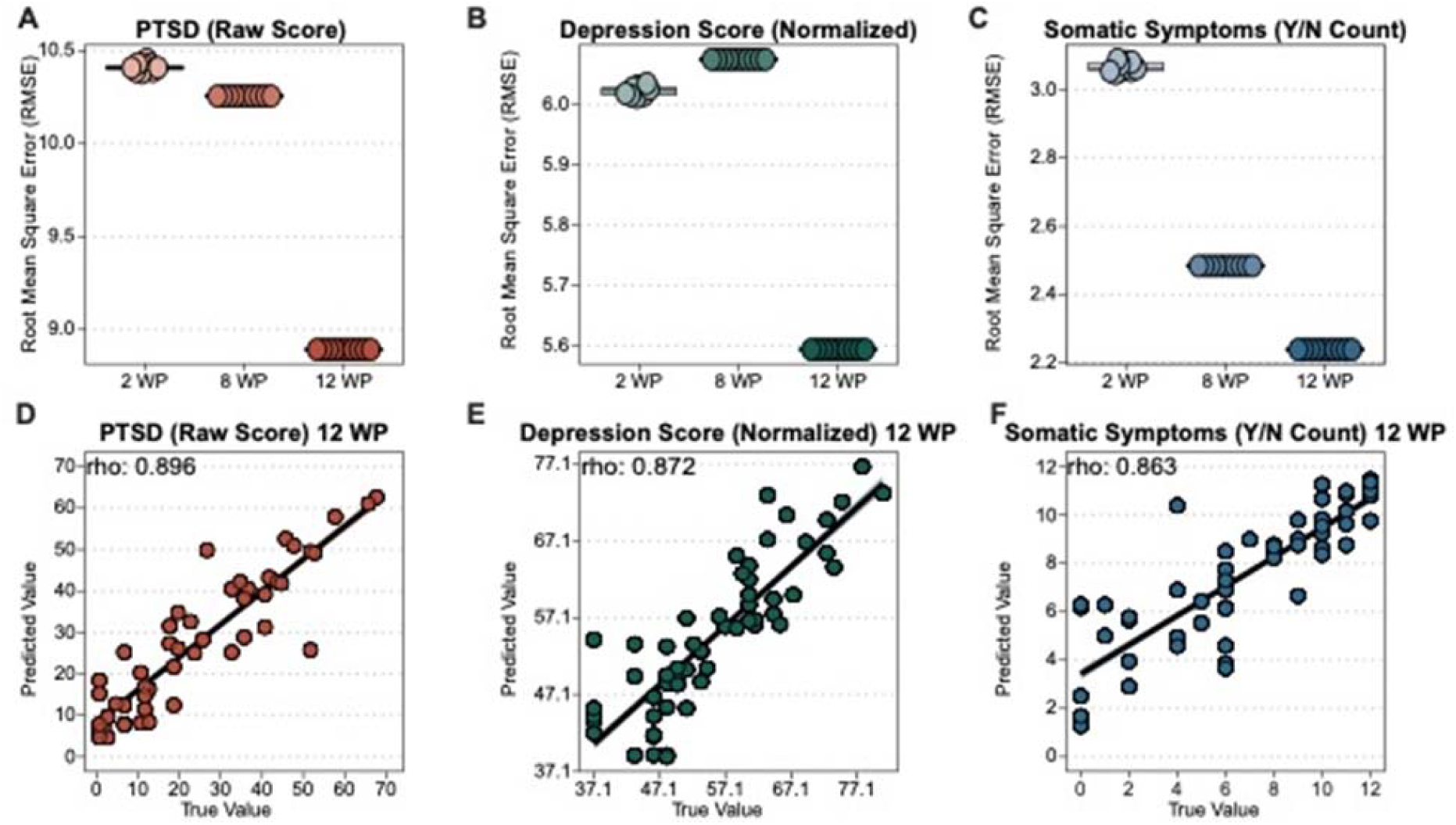
RMSE and correlation graphs for MERF models trained with microbial metabolic pathway data.

**Supplemental Figure 3.**
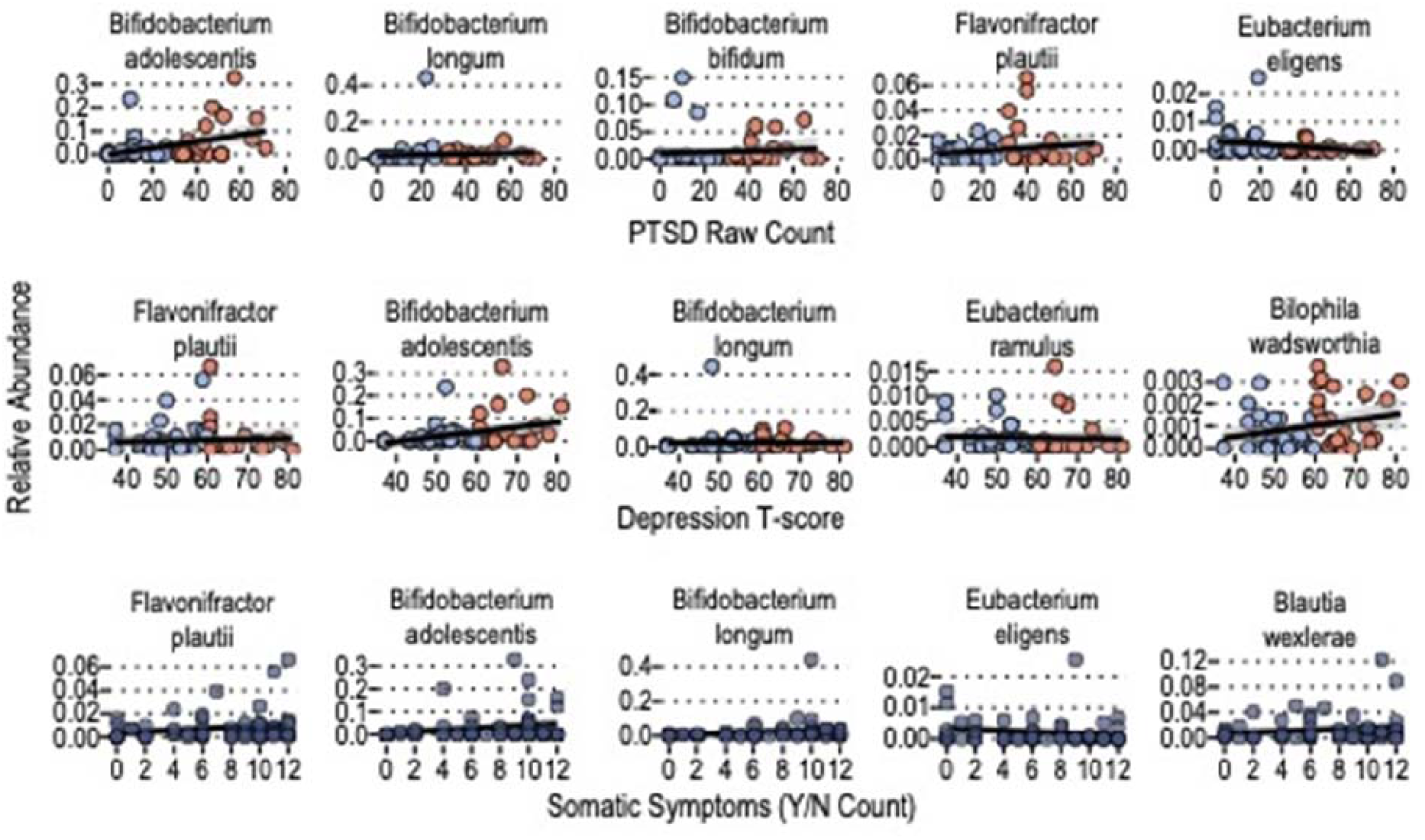
Relative abundance of MERF identified important microbes across PTSD, depressive symptoms, and somatic symptoms severity

## Notes

### Competing Interest Statement

The authors declare the following competing interests. Dr. Neylan has received research support from NIH, VA, and Rainwater Charitable Foundation, and consulting income from Boehringer Ingelheim and Jazz Pharmaceuticals. Dr. Clifford has received research funding from the NSF, NIH and LifeBell AI, and unrestricted donations from AliveCor Inc, Amazon Research, the Center for Discovery, the Gates Foundation, Google, the Gordon and Betty Moore Foundation, MathWorks, Microsoft Research, Nextsense Inc, One Mind Foundation, the Rett Research Foundation, and Samsung Research. Dr Clifford has financial interest in AliveCor Inc and Nextsense Inc. He also is the CTO of MindChild Medical and CSO of LifeBell AI and has ownership in both companies. These relationships are unconnected to the current work. Dr. Rauch reports grants from NIH during the conduct of the study; personal fees from SOBP (Society of Biological Psychiatry) paid role as secretary, other from Oxford University Press royalties, other from APP (American Psychiatric Publishing Inc.) royalties, other from VA (Veterans Administration) per diem for oversight committee, and other from Community Psychiatry/Mindpath Health paid board service, including equity outside the submitted work, other from National Association of Behavioral Healthcare for paid Board service, other from Springer Publishing royalties, and Leadership roles on Board or Council for SOBP, ADAA (Anxiety and Depression Association of America), and NNDC (National Network of Depression Centers). Dr. Sheikh has received funding from the Florida Medical Malpractice Joint Underwriters Association, Dr. Alvin E. Smith Safety of Healthcare Services Grant, the Allergan Foundation, the Jacksonville Aging Studies Center, the Substance Abuse and Mental Health Services Administration, and the Florida Blue Foundation. Dr. Jones has no competing interests related to this work, though he has been an investigator on studies funded by AstraZeneca, Vapotherm, Abbott, and Ophirex. Dr. McLean served as a consultant for Walter Reed Army Institute for Research and for Arbor Medical Innovations. Dr. Kessler was a consultant for Cambridge Health Alliance, Canandaigua VA Medical Center, Holmusk, Partners Healthcare, Inc., RallyPoint Networks, Inc., and Sage Therapeutics. He has stock options in Cerebral Inc., Mirah, PYM, and Roga Sciences. Dr. Koenen has been supported by the Robert Wood Johnson Foundation, the Kaiser Family Foundation, the Harvard Center on the Developing Child, Stanley Center for Psychiatric Research at the Broad Institute of MIT and Harvard, the National Institutes of Health, One Mind, the Anonymous Foundation, and Cohen Veterans Bioscience. She has been a paid consultant for Baker Hostetler, Discovery Vitality, and the Department of Justice. She has been a paid external reviewer for the Chan Zuckerberg Foundation, the University of Cape Town, and Capita Ireland. She has had paid speaking engagements in the last three years with the American Psychological Association, European Central Bank. Sigmund Freud University in Milan, Cambridge Health Alliance, and Coverys. She receives royalties from Guilford Press and Oxford University Press. Dr. Bucci reports funding from the National Institute of Health, the Department of Defense, and the Bill & Melinda Gates Foundation for unrelated work. Dr. Bucci is also supported by a Sponsored Research Agreement from Vedanta Biosciences, Inc, which is unrelated to the present work. 

### Funding Statement

This work was supported by R01 AG067483-01 to J.H. and partially by the CDMRP PRMP W81XWH2020013 to V.B. We would also like to thank Dr. Cindy Lai for her assistance with preparing and editing this manuscript. 
The AURORA (parent) study was supported by NIMH under U01MH110925, the US Army MRMC, One Mind, and The Mayday Fund. The content is solely responsibility of the authors and does not necessarily represent the official views of any of the funders. Data and/or research tools used in the preparation of this manuscript were obtained from the National Institute of Mental Health (NIMH) Data Archive (NDA). NDA is a collaborative informatics system created by the National Institutes of Health to provide a national resource to support and accelerate research in mental health. Dataset identifier(s): NIMH Data Archive Digital Object Identifier (DOI) 10.15154/1528593. This manuscript reflects the views of the authors and may not reflect the opinions or views of the NIH or of the Submitters submitting original data to NDA.

### Author Declarations

This study was approved by the institutional review board (protocol #17-0703) from the the University of North Carolina and approved by the IRB at the University of Massachusetts Chan Medical School,

